# An E-value-Informed Sensitivity Analysis Framework for Hybrid Controlled Trials

**DOI:** 10.64898/2026.03.05.26347653

**Authors:** Chunnan Liu, Melanie Mayer, Kimberly Lactaoen, Lizbeth F. Gomez, Gary E. Weissman, Rebecca A. Hubbard

## Abstract

Hybrid controlled trials (HCTs) incorporate real-world data into randomized controlled trials (RCTs) by augmenting the internal control arm with patients receiving the same treatment in routine care. Beyond increasing power, HCTs may improve recruitment by supporting unequal randomization ratios that increase patient access to experimental treatments. However, HCT validity is threatened by bias from unmeasured confounding due to lack of randomization of external controls, leading to outcome non-exchangeability between internal and external control patients. To address this challenge, we developed a sensitivity analysis framework to assess the robustness of HCT results to potential unmeasured confounding. We propose a tipping point analysis that adapts the E-value framework to the HCT setting where trial participation rather than treatment assignment is subject to confounding. To aid interpretation, we also introduce a data-driven benchmark representing the strength of unmeasured confounding reflected by the observed outcome non-exchangeability. We then propose an operational decision rule and evaluate its performance through simulation studies. Finally, we illustrate the approach using an asthma trial augmented by data from electronic health records. Simulation results demonstrate that our decision rule safeguards against Type I error inflation while preserving the power gains achieved by incorporating external data. In settings where moderate unmeasured confounding led to poorer outcomes for external controls, Type I error was controlled near the nominal 5% level, and power increased by 10–20% compared with analyses using RCT data alone. Our approach provides a practical, interpretable method to assess HCT robustness, supporting rigorous inference when integrating external real-world data.

## 1 Introduction

Hybrid controlled trials (HCTs) represent an innovative approach to clinical trial design in which the internal control arm of a randomized controlled trial (RCT) is augmented with external real-world data (RWD) from patients receiving the same treatment in routine care. This design is motivated by the desire to improve trial efficiency, reduce patient burden, and expedite the development of effective therapies, particularly for rare diseases or when standard-of-care treatments are minimally effective or toxic^1–4^. However, the integration of external data introduces multiple sources of potential bias, including but not limited to unmeasured confounding, baseline incomparability, outcome incomparability, and lack of concurrency^2^. In particular, unmeasured confounding arising from systematic differences in unmeasured characteristics between patients in the RCT and the external control arm poses a significant threat to the validity of HCTs, as it can lead to spurious associations or mask true treatment effects. Analysis of HCTs typically uses causal inference or Bayesian methods. Causal inference uses propensity scores to balance covariates, while Bayesian borrowing adjusts the amount of data used from RWD to account for differences between the control arms^5–17^. While there are many analysis methods for HCTs, few focus on sensitivity analysis for unmeasured confounding.

Unmeasured confounding in HCTs arises when one or more unobserved patient characteristics affect treatment assignment through differential inclusion in the RCT versus the external cohort, and also affect the outcome of interest, distorting the estimated treatment effect. The residual difference (RD), defined as the difference in the outcome between the external control and internal control arms after accounting for measured confounders, is a key indicator often used to reflect this unmeasured confounding in HCTs^18,19^. Some prior work has explored sensitivity analysis approaches for unmeasured confounding in HCTs. Yi et al. (2023)^19^ proposed a tipping point analysis for HCTs based on RD, focusing on the maximum RD that still results in statistical significance of the treatment effect. Li and Jemielita (2023)^18^ introduced a bias correction procedure for HCTs, where RD is used to estimate and partially correct for bias in the estimated effect of a treatment on a survival outcome. Though these works provide alternative sensitivity analysis approaches for unmeasured confounding, they focused on RD as an indirect measure of the magnitude of unobserved confounding as opposed to focusing directly on the impact of unmeasured confounding on the treatment effect. This can make it challenging to intuitively evaluate the magnitude of confounding and the plausibility of such a hypothetical unobserved confounder. Additionally, the existing work often focuses on one outcome type and a corresponding effect measure. Though these approaches can be generalized to other types of outcomes, the generalization process may not be straightforward. Finally, the existing work is generally technically complex, making it difficult to implement and lacking in transparency. Clear communication regarding unobserved confounding is essential for regulatory approval and for informing the clinical application of products evaluated with HCTs. Additional work is therefore needed to establish the mechanistic relationship between an unobserved confounder of the trial inclusion/outcome relationship and the magnitude of bias introduced into the treatment effect estimate. This would support a sensitivity analysis framework that is easy to implement across diverse outcome types and effect measures.

One widely recognized framework for sensitivity analysis for unobserved confounding is the E-value, proposed by Ding and VanderWeele^20,21^. The E-value quantifies the minimum strength of association that an unmeasured confounder would need to have with both the treatment and the outcome, conditional on measured covariates, to explain away an observed treatment effect. Its appeal lies in its simplicity, interpretability, and generalizability across different outcome types and effect estimates, with few assumptions required. One of the sensitivity analysis parameters for the E-value is the strength of the association between the unmeasured confounder and treatment. However, in the HCT context, unmeasured confounders do not have a direct effect on treatment but rather an indirect effect through trial inclusion, because the treatment is randomized within the trial and there is only one treatment within the external cohort. Therefore, a direct application of the E-value is not appropriate in the HCT context.

We propose a novel E-value–informed sensitivity analysis framework for HCTs that quantifies the maximum unobserved confounding of the trial inclusion–outcome relationship to which a treatment effect estimate is robust. This framework incorporates a benchmark derived from RD to represent the observed level of unmeasured confounding, and we use this benchmark to propose practical decision rules to assist researchers in assessing the robustness of their HCT findings. Our framework has several advantages: Built on the well-known E-value framework, it is easy to adopt; it also inherits the advantages of the E-value framework, including easy implementation, interpretation and applicability across outcome types and measures of effect with limited assumptions; beyond the E-value, the benchmark, defined on the same scale, provides additional context for interpreting the sensitivity analysis result. After formally introducing our framework, we evaluate its performance in a simulation study and illustrate its use in a published RCT of several asthma treatments augmented with external controls from the electronic health record of a large academic health system (Penn Medicine).

## 2 Methods

We consider a two-arm RCT consisting of an experimental treatment arm and an internal control arm, augmented by additional patients receiving the same control treatment from external RWD sources after applying eligibility criteria and accounting for measured confounders. Our sensitivity analysis framework builds on two complementary metrics: the HC-value, an adaptation of the E-value to the HCT context, and the RD-value, a benchmark derived from RD. We next formally define these quantities using notation and causal diagrams.

### 2.1 Notation

Let *A* denote a binary treatment, *Y* denote a binary outcome, *S* denote trial inclusion, ***X*** denote a vector of measured confounders, and *U* denote a categorical unmeasured confounder with levels 0, 1, …, *K*. Categorical confounders provide a more interpretable scale for confounding strength than continuous ones, though their derivations are parallel. The binary unmeasured confounder is a special case of this framework. In the explication below, all associations are expressed on the risk ratio (RR) scale. Nonetheless, this E-value-informed framework can be extended to other types of outcomes *Y* (e.g., time-to-event, non-negative count or continuous) and corresponding effect measures by computing approximate risk ratios^21^. Measured confounders ***X*** are omitted from further consideration below as we assume that appropriate adjustment for measured covariates has been performed in a preliminary analysis stage, e.g. propensity score matching. Figure 1a illustrates the assumed causal relationships among these variables.

**Figure 1.**
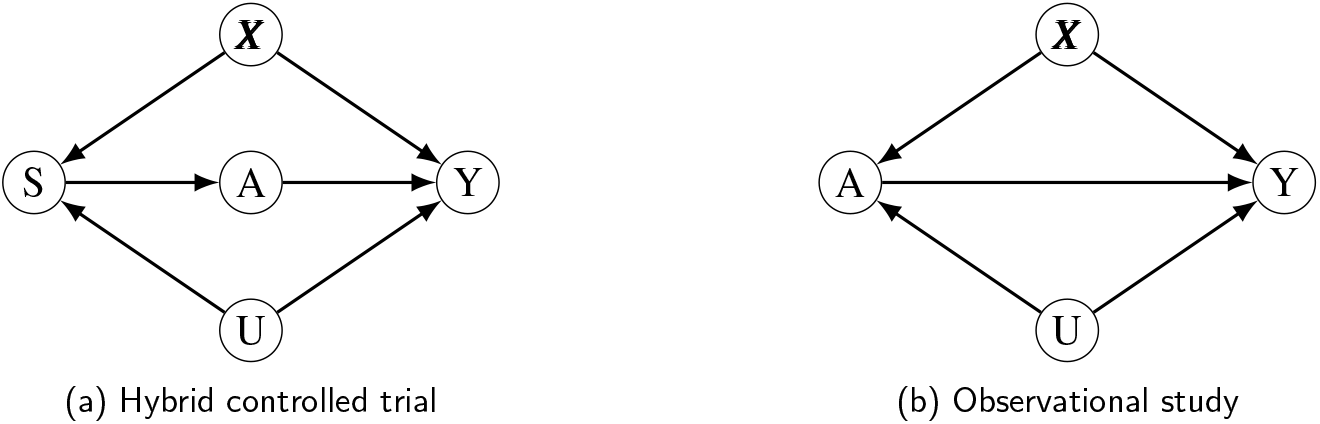
Directed acyclic graphs (DAGs) for (a) a hybrid controlled trial in which measured (***X***) and unmeasured (*U*) confounders impact trial inclusion (*S*) and outcome (*Y*), with treatment assignment (*A*) random for patients in the trial and deterministic for those in the external control arm and hence not directly affected by ***X*** or *U* ; and (b) an observational study of the effect of *A* on *Y* with measured confounders (***X***) and unmeasured confounder (*U*).

For the design of the HCT, let *α* = *P*(*A* = 1|*S* = 1)*/P*(*A* = 0|*S* = 1) denote the allocation ratio within the trial. Let *η* = *P*(*S* = 0|*A* = 0)*/P*(*S* = 1|*A* = 0) denote the relative size of the external control arm to the internal control arm, quantifying the extent of borrowing from external data. *α* and *η* may be fixed by design and hence known. Alternatively, empirical estimators for these measures are given by the sample size ratio comparing treated to internal control arms, 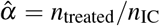, and the sample size ratio comparing the size of the external control to internal control arms, 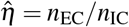.

Next, we define two sensitivity analysis parameters: *RR*_*SU*_, quantifying the imbalance of *U* between the trial and external data; and *RR*_*UY*_, quantifying the effect of *U* on the outcome *Y*. Define *RR*_*SU*_ (*k*) = *P*(*U* = *k*|*S* = 1)*/P*(*U* = *k*|*S* = 0) as the risk ratio for category *k* of *U* comparing trial participants (*S* = 1) to the external cohort (*S* = 0). Let *RR*_*SU*_ = *max*_*k*_*RR*_*SU*_ (*k*) denote the maximal risk ratio for *U* comparing *S* among all categories of *U*. Define 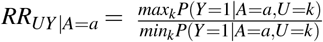 as the maximum of the effect of *U* on outcome *Y* among the treated group (*A* = 1) or control group (*A* = 0), comparing any two categories of *U*. This definition allows for interaction between *A* and *U*. Let *RR*_*UY*_ = *max*(*RR*_*UY*|*A*=1_, *RR*_*UY*|*A*=0_) denote the maximal risk ratio for *Y* comparing levels of *U*.

For treatment effects, let 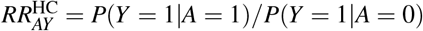 denote the HCT treatment effect. It is defined as the risk ratio for *Y* comparing the treated arm and the hybrid control arm consisting of both the internal control and external controls. Using the HCT study sample, an estimate can be obtained, 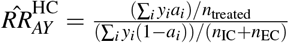. Note that the HCT treatment effect is partly attributable to the effect of *U* on *S* and *Y*. The trial treatment effect is defined as 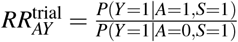, i.e. the risk ratio for *Y* comparing treatment *A* within the trial population *S* = 1. It is the true causal treatment effect because randomization of treatment assignment within the trial eliminates the effect of *U*. We can use trial data only to estimate this treatment effect, i.e. 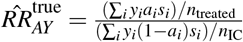.

We now introduce two important quantities that the HC-value and RD-value are built on. RD is defined as the risk ratio for the outcome comparing the internal and external control groups:

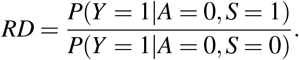

It is a direct measure of the non-exchangeability between the two control arms after accounting for measured covariates. The bias factor is defined as the ratio of the HCT treatment effect to the trial treatment effect:

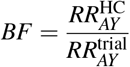

It quantifies the magnitude of bias in the treatment effect estimate induced by unmeasured confounding. In the absence of unmeasured confounding, both RD and bias factor are 1.

### 2.2 HC-value: E-value for HCT

The relationship between the bias factor and (*RR*_*SU*_, *RR*_*UY*_), is given by

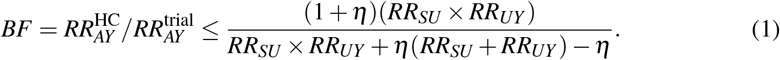

This shows the upper bound of bias induced by an unmeasured confounder with given strength of association with trial inclusion and outcome. A proof of this relationship is provided in the Appendix. This implies that,

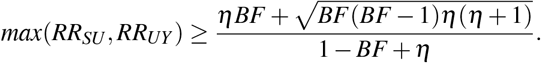

Conceptually, this bound implies that at least one of *RR*_*SU*_ or *RR*_*UY*_ must be greater than or equal to the right hand side above to induce the observed bias factor. The bound is attained if and only if *RR*_*SU*_ equals *RR*_*UY*_. We define HC-value as the right hand side when the trial treatment effect is null:

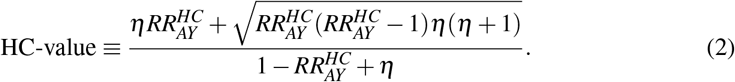

The HC-value can be interpreted as the minimum strength of association that an unmeasured confounder would need to have with either trial inclusion or the outcome to explain away the HCT treatment effect. A large HC-value implies that considerable unmeasured confounding would be needed to induce the HCT treatment effect. A small HC-value implies that even a small amount of unmeasured confounding would induce the HCT treatment effect. Thus, a small HC-value suggests that the HCT result is not robust to the potential effect of an unmeasured confounder. Operationally, we can use the HCT data to estimate the HCT treatment effect. We then plug our estimate into formula 2 to compute the HC-value of the estimate effect to obtain a quantification of robustness of this result to unmeasured confounding.

In practice, the statistical significance of the effect may be particularly important to decisionmarkers, for instance in determining whether to move a treatment forward for further evaluation in a phase III trial. Therefore, we are also interested in the unmeasured confounding strength needed to explain away the statistical significance of the HCT treatment effect. This can be represented by the HC-value corresponding to the confidence interval (CI) limit closer to the null (the minimal strength of unmeasured confounding needed to shift the CI to include the null). If the CI already includes the null, then the HC-value for the CI is 1 (no confounding is needed). While the HC-value provides a single-number minimax summary of confounding strength, a more complete sensitivity analysis considers the full set of (RRsu, RRuy) pairs that satisfy the bias-factor inequality, which can be visualized as a two-dimensional sensitivity region. An example in which we plot all values of (*RR*_*SU*_, *RR*_*UY*_) satisfying the inequality 1 is provided in the data analysis section.

### 2.3 RD-value: Residual Difference Benchmark

RD, specifcially defined in our framework as the risk ratio for outcomes comparing the external and internal control arms after accounting for measured confounders, is a key quantity in HCTs. First, it is only available in HCTs due to the availability of internal control, in contrast to fully externally controlled trials. Second, it reflects the non-exchangeability resulting from the unmeasured confounding. We can use RD to aid in the interpretation of the extent of unmeasured confounding present in the data.

We derive the relationship between RD and (*RR*_*SU*_, *RR*_*UY*_) and define RD-value based on this relationship as

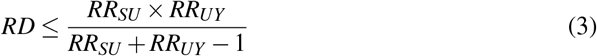

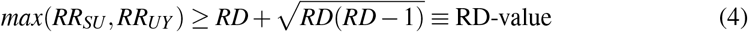

The proof is provided in the Appendix. The RD-value is interpreted as the minimum strength of association that an unmeasured confounder would need to have with either trial inclusion or the outcome to induce a given RD. Using data from the internal and external controls, we can estimate 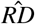 and plug it into the formula 4 to compute the RD-value. Similar to HC-value, one can also plot all values of (*RR*_*SU*_, *RR*_*UY*_) satisfying the inequality 3.

### 2.4 Sensitivity Analysis for HCT

Although the HC-value and RD-value share the same scale as max(*RR*_*SU*_, *RR*_*UY*_), they have different interpretations and offer complementary perspectives for sensitivity analysis. Intuitively, the HC-value captures the “tolerable” strength of unmeasured confounding indicative of a non-null trial treatment effect (or a statistically significant trial treatment effect), whereas the RD-value, used as a benchmark, captures the strength of unmeasured confounding implied by RD.

By definition of the bias factor, the HCT treatment effect equals the product of the bias factor induced by unmeasured confounding and the trial treatment effect. In the spirit of a proof by contradiction, we first assume that the trial treatment effect is null and attribute the entire HCT treatment effect to unmeasured confounding. We then assess the plausibility of this scenario by examining the strength of unmeasured confounding required to induce the observed HCT treatment effect. If the required magnitude of confounding is very large, it becomes implausible that the effect is entirely due to confounding, suggesting that the results are likely robust to unmeasured confounding and that a non-null trial treatment effect exists.

For RD, when conditioning on *A* = 0, there is no direct effect of *S* on *Y* in the directed acyclic graph (DAG) (Figure 1a). *S* is associated with *Y* only through *U*. Thus, RD is fully attributable to unmeasured confounding. We therefore use the magnitude of confounding associated with RD as a benchmark to provide additional context when assessing the plausibility of an unmeasured confounder with the magnitude represented by the HC-value.

In summary, the general idea behind our sensitivity analysis framework is that if the RD-value is much smaller than the HC-value, the findings can be regarded as likely to be robust to unmeasured confounding. We now formally propose a practical decision rule to assess robustness of HCT results to unmeasured confounders that affect both trial inclusion and the outcome, given the estimated HCT treatment effect 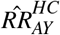 and residual difference 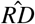.

1. Assess the statistical significance of the HCT treatment effect. If it is significant, proceed with the sensitivity analysis; otherwise, do not conduct the sensitivity analysis.
2. Compute the HC-value for both the point estimate 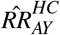 and its confidence interval limit closer to the null according to equation 2.
3. Compute the RD-value for the estimated residual difference 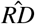 using equation 4.
4. Reject the null hypothesis if RD-value is smaller than the HC-value. Otherwise, do not reject the null even though the HCT treatment effect itself is significant. This assessment can be made either based on the HC-value for the point estimate or the confidence interval limit closer to the null, depending on whether the researcher is primarily interested in the effect estimate or its statistical significance.
5. (Optional) Plot the pairs of (*RR*_*SU*_, *RR*_*UY*_) needed to explain away the estimated HCT treatment effect 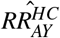 (or statistical significance) and inducing the observed residual difference 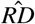 using the inequalities 1 and 3, respectively.

Note that the definitions of *RR*_*UY*_ and *RR*_*SU*_ as the maximal risk ratios lead to RD and bias factors greater than one, as well as an HCT treatment effect greater than 1 when we assume the trial treatment effect is 1. In real world application, 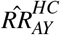 or 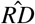 may be less than one. In that case, one first takes the inverse of these quantities before applying the formulas. Equivalently, one may add this step before HC-value and RD-value calculation:

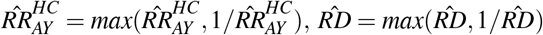

### 2.5 How does HC-value differ from the E-value?

We do not recommend using the E-value directly in the HCT context. In the standard E-value formula, one of the sensitivity analysis parameters is the strength of the association between *U* and *A*, i.e. *RR*_*AU*_. However, in the HCT context, there is no direct effect of *U* on *A* but only through *S*. The DAG in an observational study suitable for an E-value-based sensitivity analysis is shown in Figure 1b and can be compared with Figure 1a, the DAG of an HCT. Specifically, within the trial population (*S* = 1), the treatment is randomized and thus not associated with any other variable; within the external cohort (*S* = 0), patients are deterministically treated only with the standard of care control treatment. Therefore, *U* impacts *A* only through *S*.

One may still compute the E-value defined in terms of *RR*_*AU*_, the composite effect of U on A through S. However, this E-value has limited practical utility. *RR*_*AU*_ does not have a clear real-world meaning, so it is hard to assess whether its magnitude is reasonable. It also cannot be directly compared with the RD-value which is defined in terms of *RR*_*SU*_. Although *RR*_*AU*_ can be written as a function of *RR*_*SU*_ and *η* (see relationship and proof given in the Appendix), the HC-value provides a more direct and interpretable way to assess the robustness of HCT results and can be directly compared with the RD-value on the same scale. Nevertheless, the E-value remains appropriate and can still be used without modification in external controlled trials that do not include an internal control arm.

## 3 Simulation Study

We conducted a simulation study to evaluate the performance (Type I error and power) of our proposed sensitivity analysis framework and associated decision rules. We specifically examined two decision rules for rejecting the null hypothesis in the presence of a statistically significant HCT result, along with two benchmark comparator strategies:

1. Effect estimate: reject the null if the HCT result is statistically significant and the RD-value is less than the HC-value for the point estimate.
2. Statistical significance: reject the null if the HCT result is statistically significant and the RD-value is less than the HC-value for the confidence interval limit closer to the null.
3. HCT: pool the external and internal controls as the comparator, and reject the null if the treatment effect estimate is statistically significant.
4. Trial only: use only the RCT data, and reject the null if the treatment effect estimate is statistically significant.

The first rule assesses whether the strength of unmeasured confounding indicated by RD is sufficient to explain away the HCT effect estimate, while the second rule assesses whether it is sufficient to explain away statistical significance. Consequently, the second rule is expected to be more conservative, i.e., less likely to reject the null. For comparison, we also evaluated decision rules based solely on statistical significance for the primary HCT analysis and an analysis using only the trial data. The HCT decision rule does not account for unmeasured confounding and may show inflated Type I error, while the trial-only decision rule eliminates this bias at the cost of reduced power due to the exclusion of external control data.

### 3.1 Data-Generating Mechanism

Data were simulated for trial inclusion *S*, treatment assignment *A*, a binary outcome *Y* and a binary unmeasured confounder *U*. We simulated data for an HCT with 300 trial participants, randomized 2:1 to the treated and internal control arms, augmented by an external control arm. We varied the following parameters:

1. Strength of unmeasured confounding. We varied the degree of outcome non-exchangeability through the association between trial inclusion and the unmeasured confounder, *RR*_*SU*_. It was varied across three levels: 1.5 (mild), 2 (moderate), and 3 (strong confounding). We assumed that *U* was more prevalent among the trial participants, so *RR*_*SU*_ was defined as 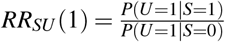. To ensure that the trial-only analysis served as a stable reference across scenarios, we fixed the prevalence of *U* at *P*(*U* = 1|*S* = 1) = 0.5 and the confounder-outcome association at *RR*_*UY*_ = 2.
2. Extent of information borrowing from RWD. The size of the external control arm was set to be 1–5 times that of the internal control arm (*η* = 1, 2, …, 5).
3. Whether external controls had better or poorer outcome than internal controls. Assuming a favorable outcome (*Y* = 1) and no interaction between *A* and *U* on *Y*, we considered two scenarios depending on the direction of the unmeasured confounding: external controls had poorer outcomes than internal controls (defined by 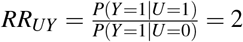) or better outcomes (defined by 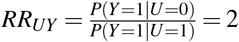). The baseline outcome prevalence among untreated individuals at the reference level of the unmeasured confounder (*P*(*Y* = 1|*A* = 0,*U* = 0)) was set to 0.2. Finally, we set the treatment effect (*RR*_*AY*_) to 1.5 for power assessment and 1.0 for Type I error assessment.

### 3.2 Performance Assessment

Each simulation scenario was repeated 5000 times. In each replication, RD and treatment effect as risk ratios were estimated using a log-binomial regression model. We assessed the statistical significance of the treatment effect at level of 0.05. We then computed the RD-value and the HC-value for both the point estimate and the confidence interval limit closer to the null. Finally, we applied the two decision rules of rejecting the null hypothesis, as well as the HCT primary analysis and the analysis using trial data only. The performance of these rules was quantified by their Type I error and statistical power, calculated as the proportion of rejections across replicates under the null and alternative hypotheses, respectively.

### 3.3 Results

Table 1 and Supplementary Table S1 summarize the distributions of key estimated quantities from the simulation, illustrating the performance of the HCT and our sensitivity metrics under various scenarios. As expected, the trial-only analysis provided an unbiased estimate of the trial treatment effect but with wider confidence intervals than the HCT analysis. Conversely, the HCT estimates were more precise but susceptible to bias, which grew in magnitude as the strength of the unmeasured confounding (*RR*_*SU*_) increased. This trend in bias was directly mirrored by RD, which deviated further from the null with stronger confounding.

**Table 1:** Simulation results varying levels of treatment effect and unmeasured confounding. Reported estimates are medians across 5000 simulation replications and empirical 95% quantiles.

| Confounding | No Treatment Effect ($RR=1$) | | | Positive Treatment Effect ($RR = 1.5$) | | |
| --- | --- | --- | --- | --- | --- | --- |
|  | Mild | Moderate | Strong | Mild | Moderate | Strong |
| Trial only result | 1.00 (0.71, 1.52) | 1.00 (0.71, 1.48) | 1.00 (0.70, 1.45) | 1.50 (1.10, 2.18) | 1.52 (1.09, 2.21) | 1.50 (1.10, 2.19) |
| RD | 1.12 (0.76, 1.52) | 1.20 (0.82, 1.64) | 1.28 (0.88, 1.77) | 1.12 (0.78, 1.52) | 1.19 (0.82, 1.64) | 1.28 (0.88, 1.76) |
| HCT result | 1.10 (0.85, 1.40) | 1.16 (0.89, 1.48) | 1.23 (0.94, 1.56) | 1.65 (1.35, 2.02) | 1.74 (1.41, 2.16) | 1.84 (1.49, 2.26) |
| RD-value | 1.60 (1.09, 2.44) | 1.72 (1.10, 2.66) | 1.89 (1.14, 2.94) | 1.59 (1.09, 2.45) | 1.71 (1.10, 2.69) | 1.89 (1.14, 2.93) |
| HC-value | 1.55 (1.09, 2.42) | 1.70 (1.10, 2.66) | 1.89 (1.14, 2.92) | 3.19 (2.25, 4.50) | 3.51 (2.45, 5.09) | 3.84 (2.69, 5.48) |
| HC-value of CI limit | 1.00 (1.00, 1.53) | 1.00 (1.00, 1.71) | 1.00 (1.00, 1.90) | 2.25 (1.46, 3.22) | 2.47 (1.62, 3.61) | 2.70 (1.80, 3.85) |

The central concept that motivates our framework is the relationship between the RD-value and the HC-value. When a trial treatment effect was present, the median HC-value was substantially larger than the median RD-value, suggesting the observed confounding was insufficient to nullify the treatment effect. These patterns held for scenarios where the external control was prognostically worse (Table 1) and better (Supplementary Table S1). Under a null treatment effect, the summary statistics show subtle differences between the RD-value and HC-value, therefore, we computed the Type I error rate and power to make pairwise comparisons as shown below.

Figure 2 illustrates the operating characteristics of the four decision rules under the scenario where the external control had worse outcomes, a common situation in practice (see Supplementary Figure S1 for the scenario when the outcome is better in external controls). In general, the analysis using trial data only had stable performance regardless of varied confounding strength. For the other three decision rules that incorporate external control, Type I error rates inflated with increasing unmeasured confounding strength (*RR*_*SU*_), and confounding inflated the treatment effect estimate away from the null, thereby increasing power. Both decision rules derived from our sensitivity analysis framework demonstrated desirable properties. They maintained lower rates of Type I error compared to the HCT analysis while achieving higher power than the analysis using only trial data across all confounding levels. The decision rule based on the effect estimate had a modest reduction in Type I error relative to the HCT. It is important to note that this reduction in Type I error, though modest, is an additional safeguard against unmeasured confounding, building upon the HCT’s initial adjustment for measured covariates. In contrast, the decision rule based on the confidence limit was more successful at controlling Type I error, yielding a Type I error rate similar to the analysis using only trial data while preserving a power gain. This decision rule strictly controls Type I error by requiring a stricter criterion: the RD-value must be smaller than the HC-value’s lower confidence limit, rather than the point estimate. Simultaneously, it preserves power through the increased sample size and the upward bias due to poorer outcomes in the external data. Results under varying *η* are provided in Supplementary Figure S2.

**Figure 2.**
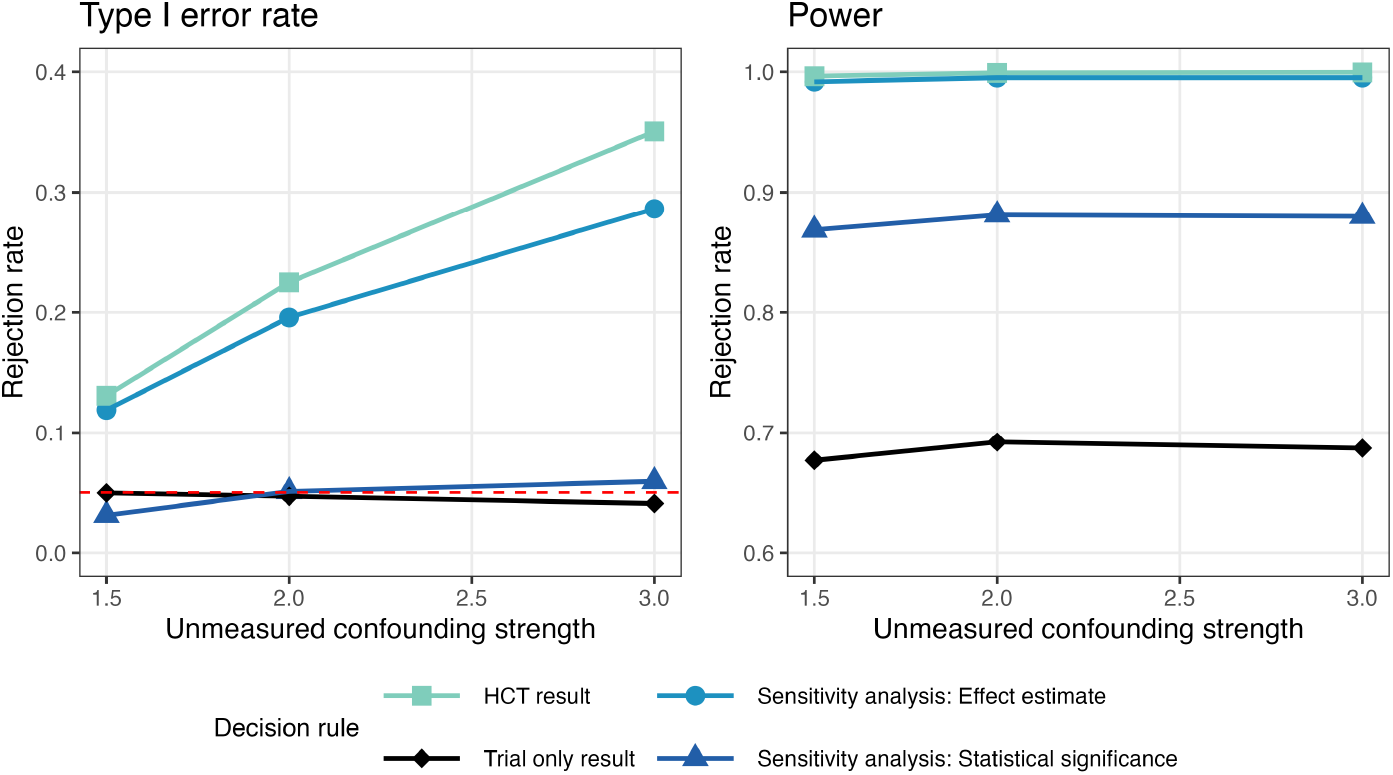
Type I error (left) and power (right) of decision rules for rejecting the null hypothesis varying unmeasured confounding strength (*RR*_*SU*_ = 1.5, 2, 3; *RR*_*UY*_ = 2) when external controls had poorer outcomes than internal controls and external control sample size was 5 times the internal control sample size (*η* = 5).

## 4 Analysis of Asthma HCT

### 4.1 Asthma Data

We applied our proposed sensitivity analysis framework to assess the robustness of the findings from an HCT for asthma treatment constructed by integrating data from a published RCT, IRIDIUM^22^, with an external control arm consisting of electronic health record (EHR) data obtained from Penn Medicine. Asthma is a chronic inflammatory lung disease characterized by episodes of airway obstruction. It represents a significant global health burden, affecting approximately 300 million individuals worldwide. People with asthma can have severe exacerbations, which require urgent treatment and can cause death^23^.

EHR data for external controls come from Penn Medicine, a large academic health care system serving the greater Philadelphia area. These data included information on prescriptions, diagnoses, and medical encounters for adult patients with asthma who received care within Penn Medicine between January 1, 2017, and February 29, 2024.

The IRIDIUM study was a randomized controlled phase III trial that assessed the efficacy of different combinations and doses of inhaled medications taken daily to control asthma. The treatment arms included mometasone furoate and indacaterol acetate (MF-IND), mometasone furoate, indacaterol acetate, and glycopyrronium bromide (MF–IND–GLY) or fluticas-one–salmeterol (FLU–SAL) in patients with inadequately controlled asthma. Patients aged 18 to 75 years with symptomatic asthma despite treatment with medium-dose or high-dose combination of an inhaled corticosteroid and a long-acting beta_2_-agonist (ICS–LABA) and at least one exacerbation in the previous year were included. Enrolled patients were randomly assigned (1:1:1:1:1) to receive medium-dose or high-dose MF–IND–GLY or MF–IND or high-dose FLU–SAL. For our analysis, the endpoint of interest was the occurrence of any severe asthma exacerbation by week 52. Severe exacerbation was defined as an aggravation of asthma symptoms that required systemic corticosteroids or an emergency room visit, hospitalization, or death due to asthma.

### 4.2 Analysis

The design and analysis of the asthma HCT adhered to FDA guidelines^3^ and, to the extent possible, the six conditions described by Pocock^1^. Specifically, selection of the real-world control arm followed the IRIDIUM trial protocol, including baseline eligibility criteria, treatment definition and window, and outcome definition (See Supplementary Table S2). We conducted two HCTs, one in which the experimental treatment arm was the medium-dose MF-IND arm and one in which the experimental treatment arm received high-dose MF-IND-GLY. In both HCTs, internal and external controls received FLU-SAL.

Entropy balancing weights were used to balance the observed covariate distribution between internal and external controls by weighting the external control data^24^. Conventional propensity score-based methods were not feasible due to lack of availability of individual patient-level data for the IRIDIUM trial. In contrast, entropy balancing serves a similar purpose but can be carried out using only summary statistics for the trial data, i.e. moments^25^, and has been previously applied in the hybrid control context^26^. The covariates to be balanced were selected based on Table 1 in the published IRIDIUM results, as well as the availability of corresponding variables in the EHR data. These included age, sex, number of severe exacerbations in the 12 months prior to study entry, and smoking status. For the optimization parameters, uniform base weights were chosen, and the weights were normalized such that the sum of the weights for the weighted external control matched the number of observations in the internal control, providing an effective sample size for external controls equal to the internal control sample size.

A weighted log-binomial regression model was fit to estimate the risk ratio for having a severe exacerbation comparing the experimental treatment arm and the hybrid control arm (pooling internal controls and weighted external controls with entropy balancing weight). Both point estimates and 95% confidence intervals are reported.

Finally, our sensitivity analysis was applied to assess the robustness of the treatment effect estimate to unmeasured confounding. RD was also estimated using a weighted log-binomial regression model with entropy balancing weights to account for observed confounders. Subsequently, RD-value and HC-value were calculated based on the estimated RD and treatment effects, and their 95% confidence intervals. For reference, the treatment effect estimated using only the trial data is also reported.

### 4.3 Results

Entropy balancing effectively balanced the covariate distribution between internal control and weighted external control arms in terms of mean and variance (Table 2). The entropy balancing weights ranged from 0.23 to 2.58, indicating the absence of extreme weights.

**Table 2:** Characteristics of internal (IRIDIUM RCT) and external control (Penn Medicine EHR) patients in asthma treatment HCT.

|  | Internal control | External control |  |
| --- | --- | --- | --- |
|  | N = 618 | Unweighted (N = 484) | Weighted (ESS = 618) |
| **Age, mean (SD)** | 52.9 (12.23) | 51.0 (13.51) | 52.9 (12.23) |
| **Sex, N (%)** |  |  |  |
| Male | 201 (32.52) | 117 (24.17) | 201 (32.52) |
| Female | 417 (67.48) | 367 (75.83) | 417 (67.48) |
| **Baseline asthma exacerbations, N (%)** |  |  |  |
| 0 | 12 (1.94) | 12 (2.48) | 12 (1.94) |
| 1 | 496 (80.26) | 335 (69.21) | 496 (80.26) |
| 2 | 94 (15.21) | 111 (22.93) | 94 (15.21) |
| 3 | 16 (2.59) | 26 (5.37) | 16 (2.59) |
| **Smoking status, N (%)** |  |  |  |
| Never | 492 (79.61) | 343 (70.87) | 492 (79.61) |
| Ever | 126 (20.39) | 141 (29.13) | 126 (20.39) |

**Abbreviations:** ESS = effective sample size after weighting. Baseline asthma exacerbations = Number of asthma exacerbations in the 12 months before the study.

For illustrative purposes, our analysis focused on the least and most effective treatments: medium-dose MF-IND (27.3% experiencing exacerbation) and high-dose MF-IND-GLY (21.8% experiencing exacerbation)^22^. In the control arms receiving high-dose FLU-SAL, the proportion of patients with exacerbation were 29.7% in the internal control arm, 41.3% in the external control arm before weighting, and 37.7% in the weighted external control arm. Despite adjustment for measured confounders, a difference of 8% between the internal control and the weighted external control arms suggests some degree of non-exchangeability, which can be attributed to unmeasured confounding. This unmeasured confounding arises because certain potential confounders were not available in our EHR data, including but not limited to the Asthma Control Questionnaire-7 (ACQ-7) and forced expiratory volume in one second (FEV1). Consequently, sensitivity analysis is crucial to evaluate the robustness of the treatment effect estimate to such unmeasured confounding.

Table 3 shows the primary analysis and sensitivity analysis results for the two experimental treatments in the HCT. The estimated HCT treatment effects were RR = 0.81 (95% CI: 0.70, 0.94) for medium-dose MF-IND and RR = 0.65 (0.54, 0.76) for high-dose MF-IND-GLY. Based on the HCT results, both treatments demonstrated statistically significant effects compared to the hybrid control arm. However, medium-dose MF-IND did not show a significant treatment effect when analyzed using only the trial data (0.92, 95% CI: 0.77, 1.10). This discrepancy could be attributed to two potential scenarios. First, the trial-only estimate is not significant due to lack of power, and the HCT estimate achieves significance due to increased sample size. Second, medium-dose MF-IND has no effect on asthma exacerbation, but the unmeasured confounding introduced by the weighted external control data led to a false-positive conclusion. The first scenario highlights the utility of HCTs, while the second represents a primary concern in their application.

**Table 3:** Primary analysis and sensitivity metrics for hybrid controlled trial of two experimental treatments for asthma (medium-dose mometasone furoate and indacaterol acetate (MF–IND) and high-dose mometasone furoate, indacaterol acetate, and glycopyrronium bromide (MF–IND–GLY)) compared to control therapy of fluticasone–salmeterol (FLU–SAL).

|  | Medium-dose MF-IND | High-dose MF-IND-GLY |
| --- | --- | --- |
| Trial-only estimate (95% CI) | 0.92 (0.77, 1.10) | 0.73 (0.60, 0.89) |
| HCT estimate (95% CI) | 0.81 (0.70, 0.94) | 0.65 (0.54, 0.76) |
| RD (95% CI) | 0.79 (0.67, 0.92) |  |
| RD-value | 1.86 (1.39) |  |
| HC-value | 2.61 (1.52) | 6.35 (3.20) |

RD-value and HC-value are reported as the value at the point estimate, with the number in parentheses computed at the 95% CI limit closer to the null.

To avoid drawing a false-positive conclusion, we conducted the proposed sensitivity analysis to assess the plausibility of an unmeasured confounder inducing the observed HCT treatment effect using the HC-value and RD-value. RD was estimated to be 0.79 (0.67, 0.92), indicating poorer outcomes in the weighted external control arm compared to the internal control arm. The implied RD-value was 1.86. The HC-values for medium-dose MF-IND and high-dose MF-IND-GLY were 2.61 (1.52 for the confidence interval limit closer to the null) and 6.35 (3.20), respectively. Intuitively, a significant conclusion is considered more robust if the HC-value exceeds the RD-value. For medium-dose MF-IND, the RD-value (1.86) was smaller than the HC-value of the point estimate (2.61), indicating that the observed unmeasured confounding may not fully explain away the treatment effect estimate; however, it was larger than the HC-value of the confidence interval limit closer to the null, indicating that the observed unmeasured confounding may fully explain away the statistical significance of the treatment effect estimate. In contrast, for high-dose MF-IND-GLY, the RD-value is much smaller than either the HC-value of the point estimate (6.35) or the confidence interval limit closer to the null (3.20), indicating that unmeasured confounding cannot fully explain away the treatment effect nor its statistical significance. Conclusions based on these findings align with the observation that the estimate using trial data alone for high-dose MF-IND-GLY was statistically significant while the result for medium-dose MF-IND was not.

More generally, pairs of *RR*_*SU*_ and *RR*_*UY*_ values that would be required to explain away the treatment effect estimate or to induce the observed RD can also be plotted (Supplementary Figure S3).

## 5 Discussion

In this paper, we proposed a novel sensitivity analysis framework for HCTs based on two metrics, the HC-value and the RD-value. Building on the well-known E-value framework, the HC-value is the minimum strength of association that an unmeasured confounder would need to have with either trial inclusion or the outcome to fully explain the treatment effect, while the RD-value provides a data-driven benchmark reflecting the confounding magnitude implied by the observed RD between control arms. This approach inherits the interpretability, ease of implementation, and flexibility of the E-value, yet extends it by incorporating a direct assessment of the non-exchangeability between internal and external controls inherent in HCTs. We proposed a practical decision rule based on not rejecting the null hypothesis if the RD-value is larger than the HC-value of the confidence interval limit closer to the null, which indicates that the observed confounding is likely to be sufficient to fully explain away the statistical significance of the HCT treatment effect. This decision rule provides a safeguard against Type I error inflation while preserving the power benefit of the HCT design in most scenarios.

This sensitivity analysis framework can also be applied at the study design stage in several ways. First, the bias due to unmeasured confounding (bias factor) can be computed under hypothesized scenarios with different external data sample sizes and levels of possible association of the unmeasured confounder with trial inclusion and outcome. To determine realistic values for the associations of the unmeasured confounder with the outcome, one can explore previously published work for prognostic factors that are not available in the data. Second, HC-values may be computed for hypothesized effect estimates. A small HC-value may indicate that one should not proceed with an HCT but should wait until resources are available to carry out an adequately powered randomized trial or invest resources in obtaining additional confounder data.

The existing work on sensitivity analyses for unmeasured confounding in HCTs^18,19^ focuses on RD as opposed to the underlying source of bias, unmeasured confounding. Additionally, they are specific to a limited set of outcome types, primarily continuous^19^ and time-to-event^18^ endpoints. Generalization to other types may not be straightforward. Finally, prior sensitivity analysis approaches, while technically sophisticated, are challenging to implement and may lack the transparency desirable to sponsors and regulators in the drug development context.

We proposed decision rules based on several approaches to using the HC-value and RD-value and their confidence intervals. Specifically, the decision rule comparing the RD-value to the HC-value calculated from the confidence interval limit closer to the null controlled Type I error, performed as conservatively as the trial-only analysis, while preserving a substantial power advantage in the common scenario where the external control group had poorer outcomes. The utility of this rule was also demonstrated in an application to an asthma HCT. For the medium-dose dual therapy, a significant HCT result that conflicted with the non-significant trial-only analysis was found to be non-robust, highlighting the decision rule’s role in mitigating the risk of Type I error. Conversely, for the high-dose triple therapy, the rule supported the robustness of a significant HCT result that was consistent with the trial-only analysis, demonstrating how the framework allows researchers to leverage the improved statistical precision due to the HCT design.

There are additional variations on these decision rules beyond the two considered in this paper. For example, one can compute the RD-value of the confidence interval for 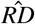 to account for the variance of the estimate. The RD-value based on the lower or upper confidence interval limit can then be compared with the HC-value. Using the RD-value of the upper limit would yield a more conservative decision rule. In particular, if one focuses on statistical significance, this choice can push the Type I error rate well below the nominal 0.05, which we view as unnecessarily conservative. For this reason, we do not recommend using the RD-value of the upper limit to compare with the HC-value of the confidence interval closer to the null. In contrast, one may use the RD-value of the lower limit to compare with HC-value of the confidence interval closer to the null if they prefer a less conservative decision rule. More generally, the decision rules can be made flexible by tuning the level of confidence. For example, using a 90% confidence interval for the HC-value rather than a 95% confidence interval relaxes the rule. This tuning parameter can be selected at the design stage, using simulations to choose the confidence level that maximizes power while maintaining the Type I error rate at 0.05.

This sensitivity analysis framework also has several limitations. First, it relies on the assumption that trial participation has no direct effect on the outcome. In practice, trial participants may receive different quality of care or have different adherence patterns compared with external controls, which can induce differences in outcomes. This assumption is strong but necessary for identification: without it, RD cannot be interpreted as a measure of unmeasured confounding between internal and external controls. Second, although our approach can be combined with frequentist methods for adjusting observed confounders, such as propensity score matching, weighting, or entropy balancing, we have not addressed how it might interact with Bayesian borrowing approaches^9,27^. Our framework currently uses point estimates and confidence intervals, but the HC-value formula could, in principle, be applied to posterior summaries obtained from Bayesian models. In that setting, the resulting HC-values would quantify sensitivity to residual bias conditional on the chosen Bayesian borrowing mechanism, remaining a causal sensitivity analysis.

As an extension of the E-value framework, our approach inherits its limitations. The E-value has been scrutinized regarding its monotonic relationship with the effect estimate and the potential for misinterpretation or automated misuse^28^. VanderWeele and colleagues have provided detailed responses and guidance for careful application; we refer readers to this exchange for a comprehensive discussion^28–30^. Beyond these general considerations, we emphasize that the RD-value in our framework does not represent the precise magnitude of unmeasured confounding in a given HCT, but rather the minimum strength of association required to induce the observed RD. Therefore, the comparison of the HC-value and RD-value does not definitively establish the existence or absence of a treatment effect, but rather serves as a sensitivity analysis assessing the robustness of the findings against the magnitude of confounding suggested by the data.

Hybrid controlled trials offer a powerful strategy to enhance trial efficiency, yet their credibility remains vulnerable to the risk of bias from unmeasured confounding. To address this fundamental challenge, we proposed a sensitivity analysis framework informed by the E-value. This provides a practical way to assess the reliability of the study findings, ensuring that the trial results are robust enough to support regulatory and clinical decision-making.

## Data Availability

Data was not collected with informed consent, and based on ethical and legal considerations, the electronic health record (EHR) data used in this work cannot be shared widely. Upon reasonable request to the authors, data may be provided.

## 6 Appendix

### 6.1 Useful lemmas

#### Lemma A.1.

*RR*_*SU*_ ≥ 1 with equality holds if and only if *S* ⊥ *U*.

*Proof of Lemma A.1*. We will prove by contradiction. Recall that 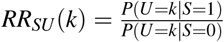 and *RR*_*SU*_ = *max*_*k*_*RR*_*SU*_ (*k*). If *RR*_*SU*_ = *max*_*k*_*RR*_*SU*_ (*k*) *<* 1, it implies *RR*_*SU*_ (*k*) *<* 1 for every *k*. We then have a contradiction:

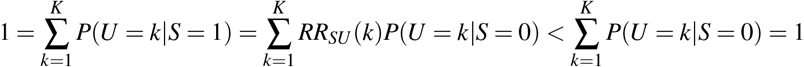

Therefore, *RR*_*SU*_ ≥ 1. Moreover, *RR*_*SU*_ = 1 implies *RR*_*SU*_ (*k*) ≤ 1 for every *k*. The only solution for 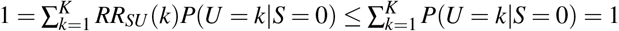 is *RR*_*SU*_ (*k*) = 1 for all *k*, i.e. *S* ⊥ *U*.

#### Lemma A.2.

When 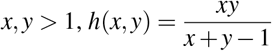 is increasing in both *x* and *y*.

*Proof of Lemma A.2*. The first partial derivative of *h*(*x, y*) with respect to *x* is

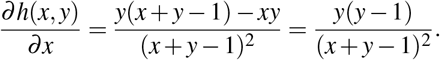

When *y >* 1, we have 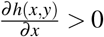, and thus *h*(*x, y*) is increasing in *x*. By symmetry, the conclusion also holds for *y* when *x >* 1.

#### Lemma A.3.

When *x, y >* 1 and 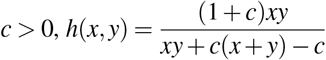 is increasing in both *x* and *y*.

#### Lemma A.4.

When 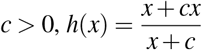 is increasing in *x*.

Proofs of Lemmas A.3 and A.4 are omitted, as they are analogous to that of Lemma A.2.

### 6.2 Relationship between the associations of the unmeasured confounder with trial inclusion and treatment

#### Proposition A.1.

*For the risk ratios for unmeasured confounder comparing trial inclusion and treatment, we have*

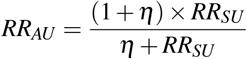

*Proof of Proposition A.1*. By definition, we have

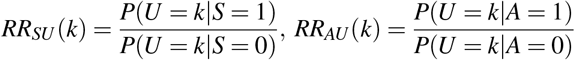

By the law of total probability, we have

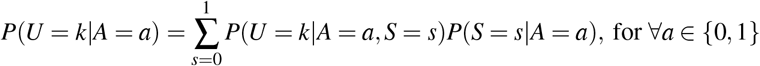

The conditional probability of *S* given *A* are:

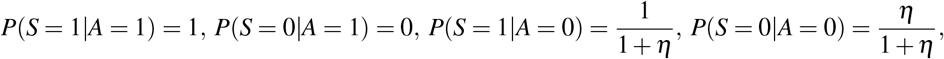

The conditional probability of unmeasured confounder are:

*P*(*U* = *k*|*A* = *a, S* = 1) = *P*(*U* = *k*|*S* = 1), ∀*a* by randomization, *P*(*U* = *k*|*A* = 0, *S* = 0) = *P*(*U* = *k*|*S* = 0)

Therefore, 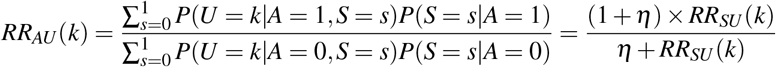

By Lemma A.4 and since *η >* 0 by definition, *RR*_*AU*_ (*k*) is maximized when *RR*_*SU*_ (*k*) = *RR*_*SU*_. Hence,

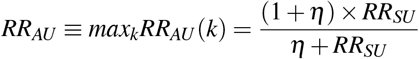

### 6.3 Derivation of residual difference and RD-value

RD is the risk ratio for outcome comparing the internal control and the external control, which reflects the non-exchangeability induced by unmeasured confounding. The proposition below quantifies this relationship and derives the formula for RD-value.

#### Proposition A.2.

*The relationship between residual difference and strength of confounding, and the inequality of RD-value are*

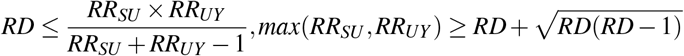

*Proof of Propositions A.2*. We will explain how to directly apply e-value for RD in the subpopulation within a hybrid controlled trial.

Since this quantity only depends on the subpopulation with *A* = 0, we would focus on that and ignore the treated arm for now. The DAG conditioning on *A* = 0 is shown in figure 3

**Figure 3.**
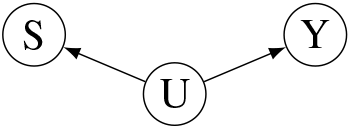
DAG representing the causal structure among patients receiving the standard of care treatment (*A* = 0), after accounting for measured confounding

This DAG would fit in a classic observational study when there is no causal effect of S on 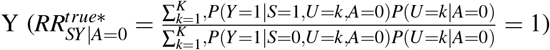, and the observed association between S and Y is fully attributed to the unmeasured confounder 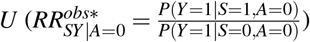. Then the e-value method can be applied treating *S* as the “treatment” here. The two sensitivity analysis parameters are 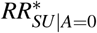 and 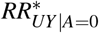. By definition:

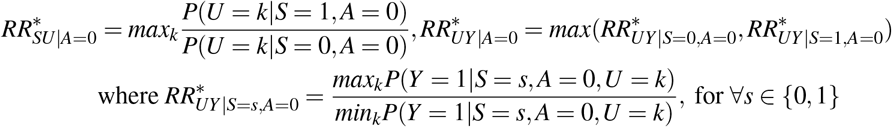

As A is randomized within S=1, and *P*(*A* = 0|*S* = 0) = 1 and *P*(*A* = 1|*S* = 0) = 0, we have:

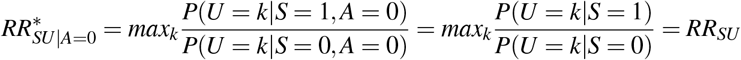

As shown in the DAG and the main text, we assumed that *Y* ⊥ *S* | (*U, A*), we have:

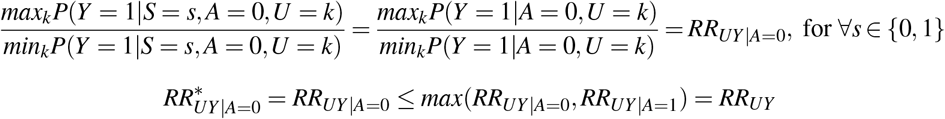

By applying e-value method, we can obtain the following results:

1. The observed treatment effect satisfy the inequality:

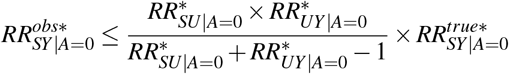

Since 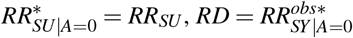, and 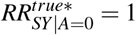, we have

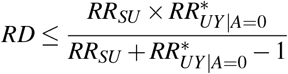

Since 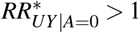 by definition, with both Lemma A.1. and A.2., and 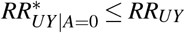:

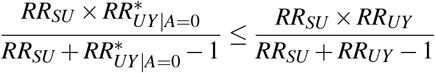

1. The two sensitivity analysis parameters satisfy the inequality:

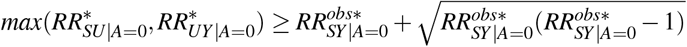

Therefore, we have:

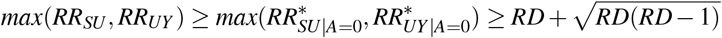

### 6.4 Derivation of bias factor and HC-value

#### Proposition A.3.

*The relationship between bias factor and residual difference is*

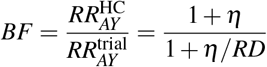

*Proof of Proposition A.3*. The treated arm was the same for both treatment effects, and the difference is in the comparators. Let 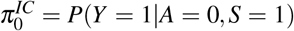 and 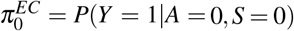. The hybrid control outcome is a weighted average of the internal control and external control outcomes:

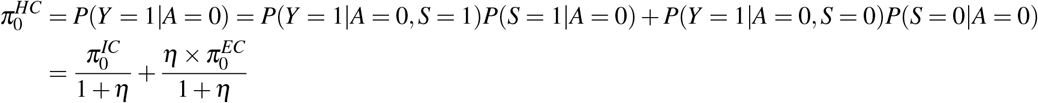

By definition, we have 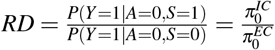. Therefore,

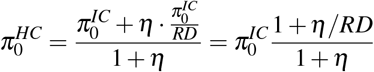

Finally we have,

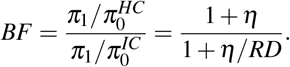

#### Proposition A.4.

*The relationship between bias factor and strength of confounding and the inequality of HC-value are*

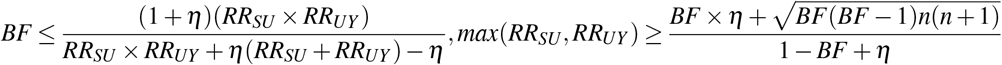

*Proof of Proposition A.4*.

From Proposition A.2, we have 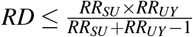.

Since *RD, η >* 0, *BF* increases as *RD* increases:

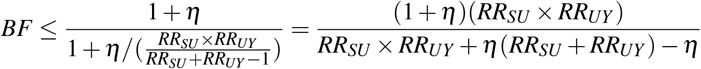

Applying Lemmas A.1 and A.3 and replacing *RR*_*SU*_ and *RR*_*UY*_ with *max*(*RR*_*SU*_, *RR*_*UY*_), we obtain the following inequality. Note that *RR*_*UY*_ *>* 1 by definition.

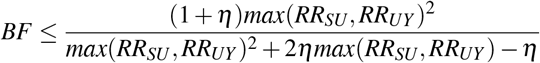

solving *max*(*RR*_*SU*_, *RR*_*UY*_) can obtain the inequality of HC-value.

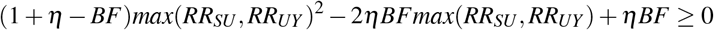

note that 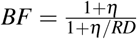 and 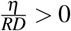, thus *BF <* 1 + *η*

## Notes

Funding: Research reported in this publication was supported by the National Heart, Lung, and Blood Institute (NHLBI), National Institutes of Health, under award number R01HL162354

### Competing Interest Statement

The authors have declared no competing interest.

### Funding Statement

Research reported in this publication was supported by the National Institutes of Health (NIH) National Heart, Lung, And Blood Institute (NHLBI) under Award Number R01HL162354

### Author Declarations

The Institutional Review Board of the University of Pennsylvania gave approval for this work.

