## Supplementary Material for "An E-value-Informed Sensitivity Analysis Framework for Hybrid Controlled Trials"

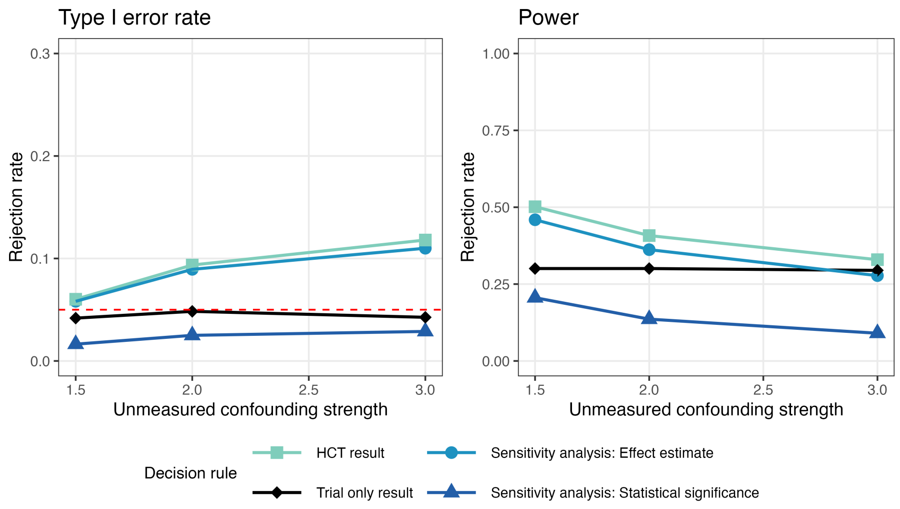


Figure S1: Type I error and power of decision rules for rejecting the null hypothesis varying unmeasured confounding strength ($RR_{SU} = 1.5, 2, 3c, RR_{UY} = 2$) when external controls had better outcomes than internal controls and external control sample size was 5 times the internal control sample size ($\eta = 5$).

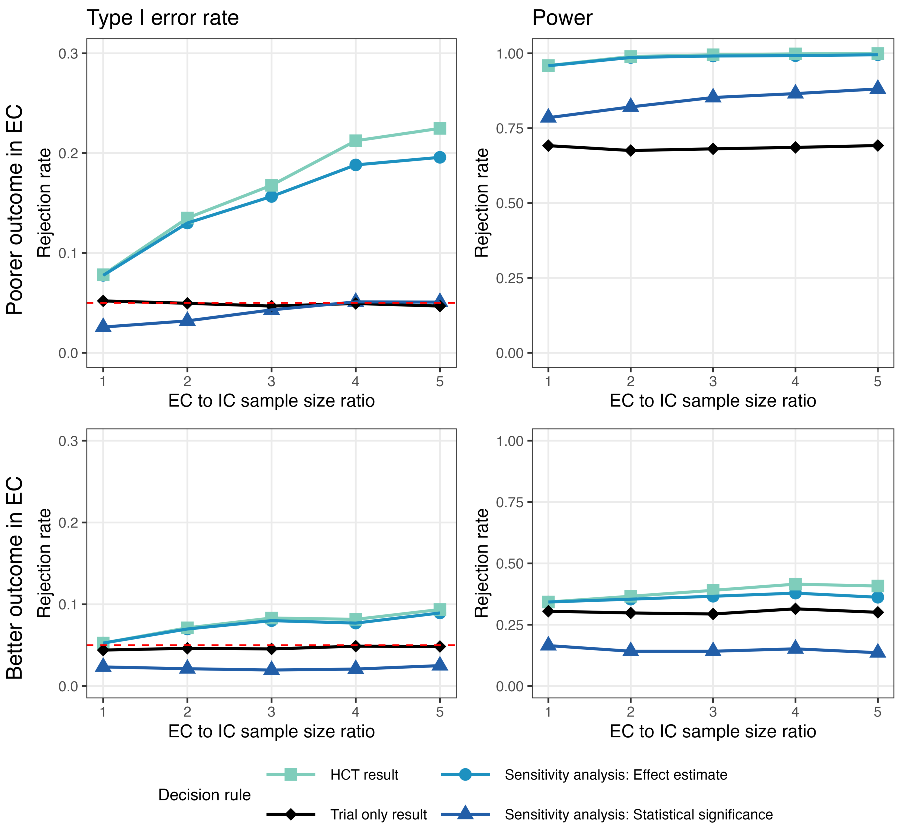


Figure S2: Type I error and power of decision rules for rejecting the null hypothesis varying external control relative sample size ($\eta = 1, 2, 3, 4, 5$) when external controls had poorer (top) or better (bottom) outcomes than internal controls, while there was strong confounding ($RR_{SU} = 3, RR_{UY} = 2$)

Table S1: Simulation results varying levels of treatment effect and unmeasured confounding when the external controls have better outcome than the internal control. Reported estimates are medians across 5000 simulation replications and empirical 95% quantiles.

| | No Treatment Effect (RR=1) | | | Had Treatment Effect (RR = 1.5) | | |
|---|---|---|---|---|---|---|
| Confounding | Mild | Moderate | Strong | Mild | Moderate | Strong |
| Trial only result | 1.00 (0.58, 1.87) | 1.00 (0.57, 1.89) | 1.00 (0.59, 1.95) | 1.50 (0.91, 2.83) | 1.51 (0.92, 2.86) | 1.50 (0.92, 2.83) |
| RD | 0.89 (0.49, 1.43) | 0.86 (0.47, 1.36) | 0.81 (0.44, 1.26) | 0.90 (0.49, 1.45) | 0.84 (0.46, 1.36) | 0.82 (0.45, 1.28) |
| HCT result | 0.91 (0.60, 1.29) | 0.87 (0.58, 1.24) | 0.85 (0.57, 1.19) | 1.37 (0.98, 1.86) | 1.31 (0.94, 1.77) | 1.27 (0.90, 1.72) |
| RD-value | 1.69 (1.12, 3.46) | 1.74 (1.12, 3.67) | 1.82 (1.12, 3.96) | 1.69 (1.12, 3.47) | 1.76 (1.12, 3.73) | 1.82 (1.12, 3.88) |
| HC-value | 1.66 (1.12, 3.26) | 1.73 (1.12, 3.47) | 1.80 (1.12, 3.55) | 2.33 (1.24, 3.93) | 2.15 (1.21, 3.62) | 2.03 (1.16, 3.43) |
| HC-value of CI limit | 1.00 (1.00, 1.40) | 1.00 (1.00, 1.58) | 1.00 (1.00, 1.62) | 1.03 (1.00, 2.35) | 1.00 (1.00, 2.18) | 1.00 (1.00, 2.06) |

Mild, moderate, strong confounding represents $RR_{SU} = 1.5, 2, 3$, respectively. This table is for the scenario in which external control outcomes were better than internal controls, specifically $RR_{UY} = 2$. HC-value of CI limit is the HC-value of the confidence interval limit closer to the null. If confidence interval includes null, then HC-value of CI limit is 1.

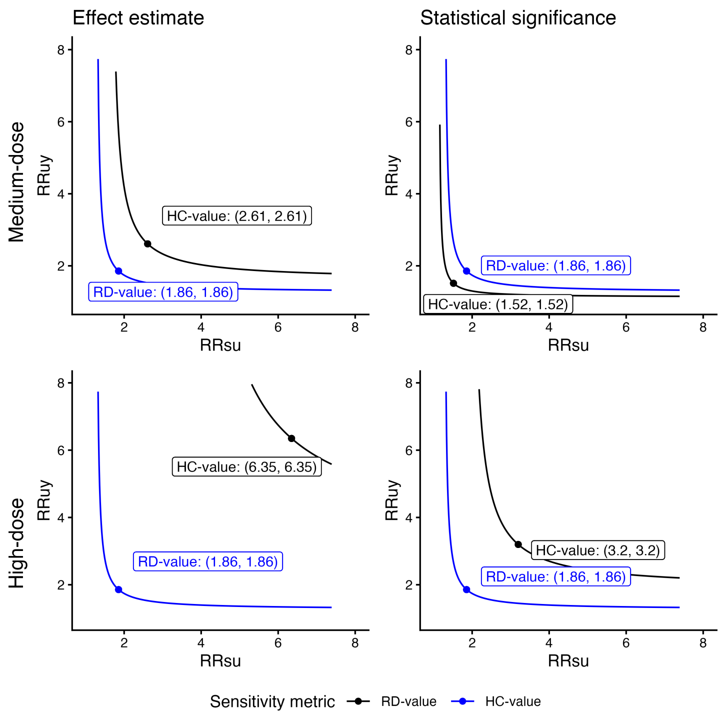


Figure S3: Joint relationship of $RR_{SU}$ and $RR_{UY}$ in asthma treatment HCTs

The top row represents the HCT with medium-dose dual therapy as the experimental treatment, and the bottom row represents the HCT with high-dose triple therapy. The left column represents the decision rule based on the effect estimate, and the right column represents the decision rule based on statistical significance. Each panel compares the unmeasured confounding strength indicated by the data (the area above the blue curve on which the RD-value lies) with the unmeasured confounding strength that is sufficient to explain away the treatment effect or statistical significance (the area above the black curve on which the HC-value lies), using the joint relationship between $RR_{SU}$ and $RR_{UY}$.

Table S2: Operational definitions and data mappings for IRIDIUM trial and EHR

| Category | Definition | Time window | EHR Dataset |
|---|---|---|---|
| **Outcome** | | | |
| Severe exacerbation | Aggravation of asthma symptoms (such as shortness of breath, cough, wheezing, or chest tightness) that requires: | After the first dose and not later than one day after the date of the last dose | diagnosis |
| | Systemic corticosteroids for at least 3 consecutive days | | prescription |
| | And a need for an emergency room visit or hospitalization owing to asthma | | encounter |
| | Or death due to asthma | | |
| **Treatment** | | | |
| HD FLU–SAL | High-dose FLU–SAL (500 $\mu$g, 50 $\mu$g) twice daily | Day 1 to day 365 | prescription |
| **Inclusion criteria** | | | |
| Age | $\geq$ 18 years old and $\leq$ 75 years | Visit 1 | baseline characteristics |
| Diagnosis of asthma | A period of at least 1 year prior to Visit 1 | 1 year prior to visit 1 | diagnosis |
| Exacerbation history | Documented history of at least one asthma exacerbation in the 12 months prior to Visit 1 | 12 months prior to visit 1 | diagnosis, prescription, encounter |
| **Exclusion criteria** | | | |
| Smoking | Smoked or inhaled tobacco products within the 6 month period prior to Visit 1, or have a smoking history of greater than 10 pack years | 6 months prior to visit 1 | baseline characteristics |
| Exacerbation | Had an asthma attack/exacerbation requiring systemic steroids or hospitalization or emergency room visit within 6 weeks of Visit 1 (Screening) | 6 weeks prior to visit 1 | diagnosis, prescription, encounter |
| Other chronic lung diseases | History of chronic lung diseases other than asthma, including (but not limited to) chronic obstructive pulmonary disease, sarcoidosis, interstitial lung disease, cystic fibrosis, clinically significant bronchiectasis, and active tuberculosis | Ever | diagnosis |
